# Epigenetic Clocks of Biological Aging and Cognitively Healthy Longevity: The Women’s Health Initiative Memory Study

**DOI:** 10.64898/2026.03.11.26348107

**Authors:** Andrea Z. LaCroix, Bowei Zhang, Steve Nguyen, Susan M. Resnick, Luigi Ferrucci, Steve Horvath, Ake T. Lu, Mark A. Espeland, Stephen R. Rapp, Kenneth Beckman, Caroline Nievergelt, Adam X. Maihofer, Linda K. McEvoy, Aladdin H. Shadyab

**Affiliations:** Herbert Wertheim School of Public Health and Human Longevity Science, University of California San Diego, La Jolla, CA, USA; Laboratory of Behavioral Neuroscience, National Institute on Aging, National Institutes of Health, Baltimore, MD, USA; Longitudinal Studies Section, Translational Gerontology Branch, National Institute on Aging, Baltimore, MD, USA; Altos Labs, Cambridge, UK; Altos Labs, San Diego, CA; Division of Gerontology and Geriatric Medicine, Department of Internal Medicine, Wake Forest University School of Medicine, Winston-Salem, NC, USA; Department of Psychiatry and Behavioral Medicine, Wake Forest University School of Medicine, Winston-Salem, NC, USA; Genomics Center, University of Minnesota, Minneapolis, MN, USA; Department of Psychiatry, School of Medicine, University of California San Diego, La Jolla, CA, USA; Veterans Affairs San Diego Healthcare System, Research Service, San Diego, CA, USA; Kaiser Permanente Washington Health Research Institute, Seattle, WA, USA; Division of Geriatrics, Gerontology, and Palliative Care, Department of Medicine, University of California San Diego, La Jolla, CA, USA

**Keywords:** Healthspan, healthy aging, brain function

## Abstract

**BACKGROUND:** Little is known about whether epigenetic age acceleration (EAA) clocks are capable of predicting exceptional longevity with or without preserved cognitive function.

**METHODS:** We examined 5844 women from the Women’s Health Initiative Memory Study. Fifteen epigenetic clocks were measured at baseline (1996-1999). Longevity outcomes were defined as: 1) survival to age 90 with preserved cognition (n=1726, 29.5%); or 2) survival to age 90 with cognitive impairment (n=956, 16.4%); vs. 3) death before age 90 (n=2611, 44.7%). Logistic regression models examined associations between the 15 clocks and survival to age 90 (vs. death before age 90), adjusting for covariates. Multinomial logistic regression models examined associations with survival to age 90 without cognitive impairment and survival to age 90 with cognitive impairment (each vs. death before age 90), also adjusting for covariates.

**FINDINGS:** Each standard deviation increase in EAA for the first-generation clocks was associated with 7%-18% reduced odds of survival to age 90 vs. earlier death. Stronger associations were observed for second- and third-generation clocks, including AgeAccelGrim2 (OR=0.66; 95% CI 0.61-0.71), PCGrimAge (OR=0.64; 95% CI 0.59-0.69), PCPhenoAge (OR=0.73; 95% CI 0.68-0.78) and DunedinPACE (OR= 0.77; 95% CI 0.72-0.82). None of the clocks was more strongly associated with survival to age 90 with preserved cognition than with survival to age 90 with cognitive impairment, relative to death before age 90.

**INTERPRETATION:** All epigenetic clocks were associated with exceptional longevity, but none were associated with cognitive healthspan. Developing clocks that can differentiate long survival with and without preserved cognitive function is critical.

## INTRODUCTION

Based on US death rates in 2023, the probability that babies born today will survive to age 90 is 30.7% for girls and 19.3% for boys.^1^ For Americans who have reached age 65, the corresponding probabilities are 35.1% for women and 24.4% for men.^1^ While survival to age 90 is no longer a rare event, the fact that so many will not survive, and others will survive with significant impairments, serves as cogent reminder of the heterogeneity^2^ in the maintenance vs. deterioration of complex underlying biological processes and real-life functional capacity in older adults. The concept of “Precision Gerontology” would be importantly advanced if we could identify biomarkers that differentiate those who will and will not achieve this advanced age with preserved function.

A growing number of epigenetic clocks have been developed to estimate biological aging as distinct from chronological age. A 2019 meta-analysis^3^ of 5 clocks estimated that each 5-year increase in epigenetic age acceleration (EAA), indicating accelerated biological aging, was associated with an 8%-15% higher risk of death independent of chronological age. EAA has also been associated^4^ with various cognitive phenotypes, including poorer cognitive test scores and dementia (Nguyen et al, 2026). However, to date, no epigenetic clock has been trained on actual longevity, defined as survival to a very old age, or healthspan, defined as survival to a very old age with preserved function (physical, cognitive, sensory or social). Thus, little is known about whether epigenetic clocks are capable of predicting longevity with or without preserved function. The objective of this study was to prospectively investigate associations of 15 epigenetic clocks with survival to age 90 with and without preserved cognition in the large Women’s Health Initiative Memory Study (WHIMS) cohort.

## METHODS

### Study Population

The Women’s Health Initiative Memory Study (WHIMS) is an ancillary study of the WHI hormone therapy trials. WHIMS was designed to investigate the effects of hormone therapy on adjudicated cognitive outcomes (no impairment, mild cognitive impairment, or probable dementia) among 7,479 women ages 65-80 years who were cognitively unimpaired at baseline in 1996-1999. Details on the WHIMS design and protocols have been published.^5^ Annual follow-up for cognitive outcomes continued through 2007. In 2008, WHIMS transitioned to annual telephone-administered cognitive assessments in the WHIMS Epidemiology of Cognitive Health Outcomes (WHIMS-ECHO) study, which followed participants for the same cognitive outcomes through 2021.^6^

This study included 5,844 WHIMS women selected for epigenetics data measurement who were eligible to survive to age 90 as of February 17, 2024 based on their birth year (Supplementary Methods and Supplementary Figure 1). DNA was extracted from blood samples collected at the WHIMS baseline visit and shipped to the University of Minnesota Genomics Center for epigenetics measurement using the Illumina Infinium MethylationEPIC v2.0 using standard protocols (Supplementary Methods).

### Epigenetic Clocks

We examined the most widely studied clocks in the literature, including AgeAccelHorvath, AgeAccelHannum, intrinsic epigenetic age acceleration (IEAA), extrinsic epigenetic age acceleration (EEAA), IEAA.Hannum, AgeAccelPheno, DNAm-based estimator of telomere length (DNAmTL), DunedinPACE, AgeAccelGrim2, and the recently developed Intrinsic Capacity^7^ (DNAmIC Age Accel). We also examined principal components (PC)-based versions of several of these clocks, including PCHorvath, PCHannum, PCPhenoAge, PCGrimAge, and PCDNAmTL. Further description of these clocks is provided in the Supplementary Methods.

For the epigenetic clocks, those for which higher values indicate accelerated biological aging relative to chronological age were expected to be associated with lower odds of survival to age 90. The three clocks reflecting slower biological aging, namely DNAmTL, its PC version PCDNAmTL, and DNAmIC, were expected to be associated with increased odds of survival to age 90. For interpretation purposes, all epigenetic clock variables were standardized with a mean of 0 and SD of 1.

### Longevity Outcomes

We first examined exceptional longevity, defined as survival to age 90 vs death before age 90. Women were followed from baseline to February 17, 2024, to ascertain survival to age 90. Deaths were verified by trained physician adjudicators using hospital records, autopsy or coroner’s reports, or death certificates. Periodic linkage to the National Death Index was performed for all participants, including those who were lost to follow-up, for verification if death certificates or medical records were unavailable.

We further examined cognitively healthy longevity, defined as follows: (1) survived to age 90 with cognitive impairment (i.e, MCI or probable dementia); (2) survived to age 90 without cognitive impairment; or (3) died before age 90 regardless of cognitive status. This approach was used in a previous WHI analysis.^8^ MCI and dementia were ascertained and adjudicated annually through December 31, 2021. The WHIMS protocol for detecting MCI and dementia is described elsewhere and in the Supplementary Methods.^5^

For women who survived to age 90, we prioritized WHIMS cognitive assessments within 3 years of the 90^th^ birth year to determine adjudicated cognitive impairment status at age 90. For those who did not have a WHIMS cognitive assessment within 3 years of the 90^th^ birth year (n=1173), we assessed cognitive impairment at age 90 using the health status update questionnaires that were administered to all WHI participants at baseline, 1-year and 3-year follow-up assessments, and then annually after 2005. This questionnaire ascertained self-reported moderate or severe memory problems or physician-diagnosed dementia or Alzheimer disease; if either of these conditions was reported, women were classified as having cognitive impairment.

### Covariates

Baseline questionnaires assessed age, race (American Indian/Alaskan Native, Asian, Native Hawaiian or other Pacific Islander, Black, White, more than once race, or unknown/not reported), ethnicity (Hispanic/Latino, not Hispanic/Latino, or unknown/not reported), education (less than high school equivalent, high school diploma or GED, some college, college graduate), smoking status (never smoked, past smoker, current smoker), treated diabetes, cardiovascular disease, cancer, and total energy expenditure from recreational physical activity (in MET-hours/week). Height and weight were measured with a stadiometer and balance beam scale, respectively, to calculate body mass index (BMI; kg/m^2^). *APOE* ε2 and ε4 carrier status, defined as presence of at least 1 ε2 or ε4 allele, respectively, was determined in women with available genome-wide genotyping data based on 2 single nucleotide variants, rs429358 and rs7412. Hormone therapy treatment arm (randomized estrogen alone, estrogen placebo, estrogen plus progestin, or estrogen plus progestin placebo) was also included as a covariate. Finally, to account for cell types known to have different DNA methylation patterns that change with age, we adjusted for white blood cell (WBC) counts, which were estimated for CD4+ T cells, CD8+ T cells, natural killer cells, B cells, monocytes, neutrophils, and granulocytes using the Identifying Optimal Libraries (IDOL) algorithm.^9^

### Statistical Analysis

Baseline characteristics were reported by quartiles of DunedinPace and AgeAccelGrim2, which were the two clocks we were most interested in based on our prior studies showing strongest associations with MCI/ADRD and plasma biomarkers of ADRD pathology.^10–12^ The differences between quartiles were tested using the Kruskal-Wallis rank sum test for continuous variables, and Pearson’s chi-square test or Fisher’s exact test for categorical variables.

To examine the associations of epigenetic age acceleration (EAA) with longevity outcomes, we conducted logistic regression and multinomial logistic regression models to estimate odds ratios (OR) and their 95% confidence intervals (CIs) for 1 standard deviation (SD) increases in EAA measures. Separate models were fit for each EAA measure. Logistic regression models assessed the association between EAA and survival to age 90 versus death before age 90 (reference category). Multinomial logistic regression models assessed the associations of EAA with cognitively healthy longevity by comparing women who survived to age 90 with and without cognitive impairment to those who died before age 90 (reference category). Minimally adjusted models were adjusted for chronological age, hormone therapy study arm, education, smoking status, race, and ethnicity. Fully adjusted models were additionally adjusted for physical activity, BMI, diabetes, cardiovascular disease, cancer, and also for white blood cell composition (CD8 T, CD4 T, natural killer, B cell, monocyte, neutrophil). Models for IEAA, EEAA, or IEAA.Hannum did not include white blood cell counts, given that IEAA is independent of white blood cell counts and EEAA tracks age-related changes in white blood cell-type composition. Missing data in covariates were imputed using multivariate imputation by chained equations with 20 imputations and 20 iterations using the “mice” package in R. The ORs for all measures were very similar whether minimally or fully adjusted so the fully adjusted models are presented.

In sensitivity analyses, we examined effect modification of EAA associations by *APOE* ε2 and *APOE* ε4 carrier status through stratification and conducting likelihood ratio tests to determine significance. APOE genotypes were only available in White women, so these analyses were restricted to that group.

The present study was designed to evaluate the magnitude of associations of 15 epigenetic clocks with long survival with and without preserved cognition. It was hypothesis driven in the sense that each clock, as a measure of biological aging and healthspan, was postulated to be associated with survival to age 90 overall and survival to age 90 with preserved cognition vs. earlier death. The study was not designed to test the statistical superiority of any one clock vs. another, thus comparisons between clocks should be considered exploratory. All analyses were conducted using R statistical software version 4.4.2.

## RESULTS

Higher chronological age, lower education, current smoking, Black race, Hispanic ethnicity, lower physical activity, higher BMI, diabetes, and cardiovascular disease were all associated with faster EAA by AgeAccelGrim2 (Table 1). These associations were largely similar for DunedinPACE (Supplemental Table 2).

**Table 1:**
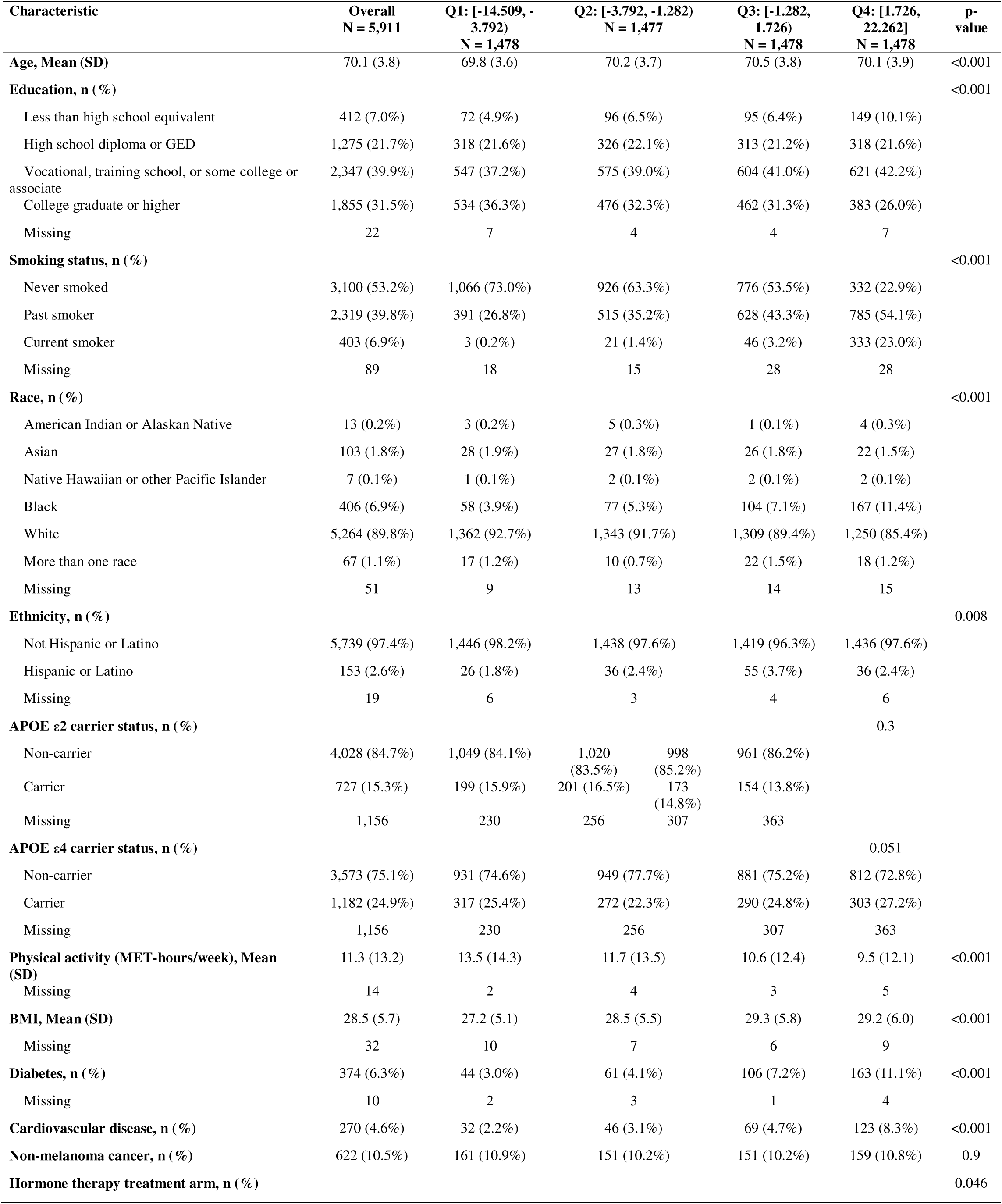
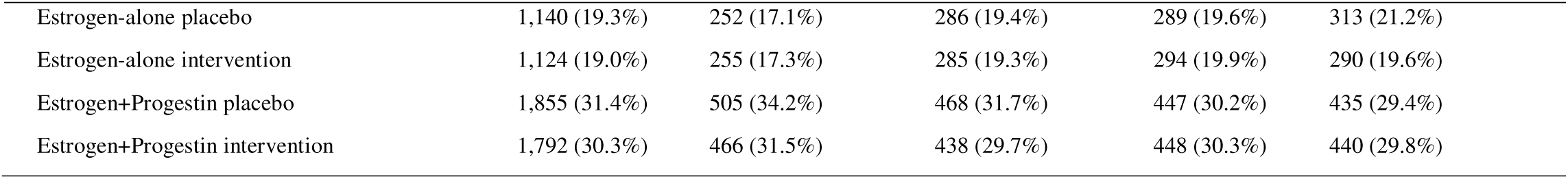
Baseline characteristics by quartiles of AgeAccelGrim2 in 5911 women enrolled in the Women’s Health Initiative Memory Study (WHIMS) who could have survived to age 90.

Of the 5,844 women eligible to survive to age 90 in the analytic study population, 3,233 (55.3%) survived, including 1,726 (29.5%) with preserved cognition and 956 (16.4%) with cognitive impairment. All 15 EAA clocks examined were associated with survival to age 90 vs. death before age 90 (Figure 1). Each standard deviation increase in EAA for the first-generation clocks (AgeAccelHorvath, AgeAccel Hannum, IEAA, EEAA, and IEAA Hannum) reduced the odds of survival to age 90 by 7%-18%. AgeAccelPheno, a second-generation clock, was also within that range of association. The PC versions of the Horvath and Hannum clocks had stronger ORs than the original versions, with ORs of 0.83 (95% CI 0.78-0.89) and 0.81 (95% CI 0.76-0.87), respectively. The strongest associations were observed for AgeAccelGrim2 (OR=0.66; 95% CI 0.61-0.71), PCGrimAge (OR=0.64; 95% CI 0.59-0.69), PCPhenoAge (OR=0.73; 95% CI 0.68-0.78) and DunedinPACE (OR= 0.77; 95% CI 0.72-0.82). Of the 3 clocks measuring slower biological aging, the strongest association observed was for PCDNAmTL, with an OR of 1.35 for survival to age 90 (95% CI 1.26-1.44). Intrinsic Aging (DNAmIC Age Accel), the newest clock examined, had a modest association with survival to age 90 (OR=1.15; 95% CI 1.09-1.23).

**Figure 1:**
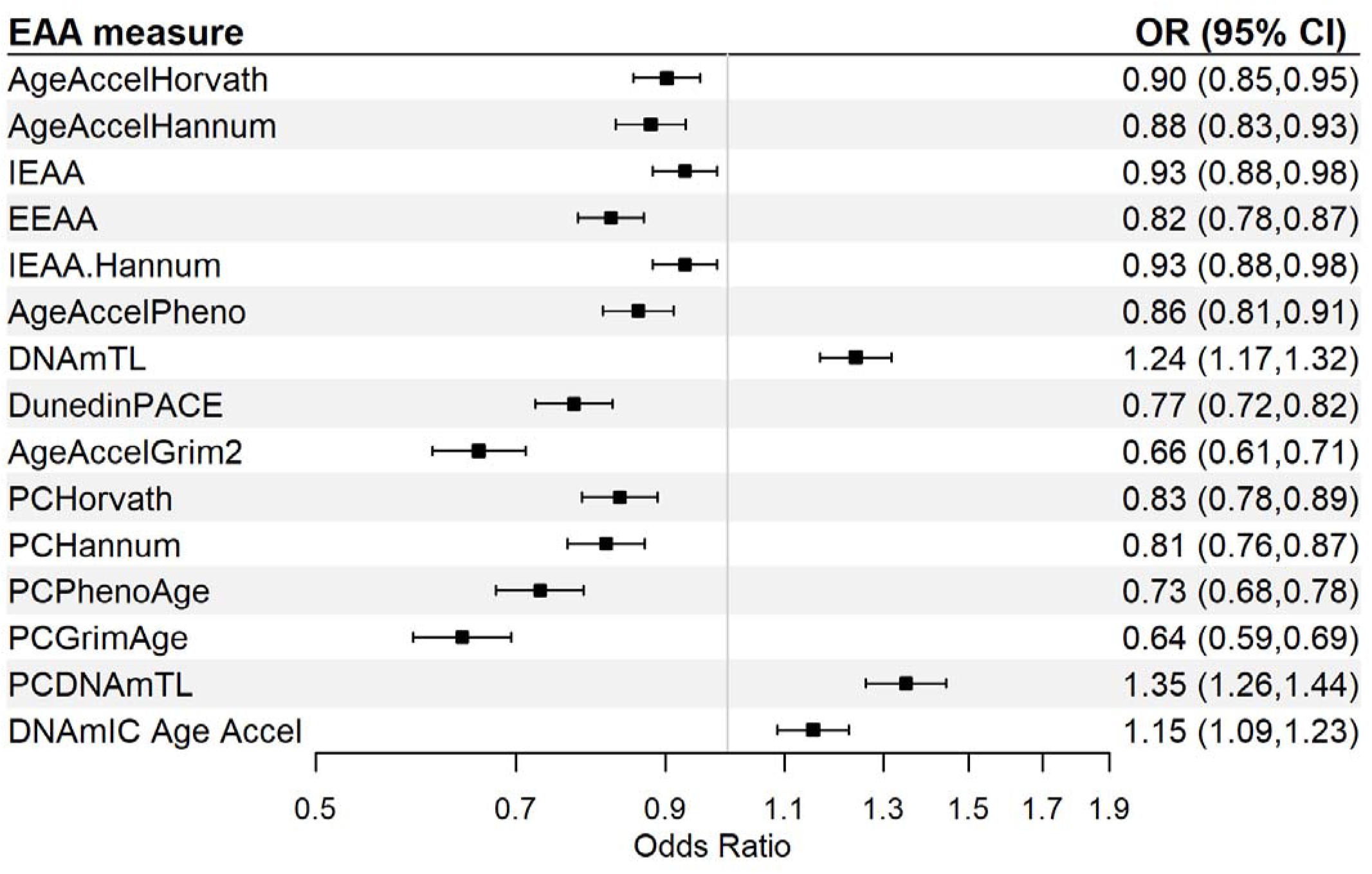
Odds ratios relating 15 epigenetic clocks to survival to age 90 in 5844 women enrolled in the Women’s Health Initiative Memory Study (WHIMS) who could have survived to age 90 1. The reference group was women who died before age 90 years (N = 2,611). 2. Odds ratios were adjusted for chronological age, race, ethnicity, hormone therapy treatment arm, education, smoking status, physical activity, body mass index, diabetes, cardiovascular disease, non-melanoma cancer, and white blood cell counts (CD8T, CD4T, natural killer cells, B cells, monocytes, and neutrophils). 3. Models with IEAA, EEAA, and IEAA.Hannum did not include white blood cell counts.

The odds of survival to age 90 with and without cognitive impairment compared to death before age 90 showed a similar pattern in the odds ratios as those described above (Figure 2). The five first-generation clocks (AgeAccelHorvath, AgeAccelHannum, IEAA, EEAA and IEAA Hannum) had ORs in the range of 0.78-0.89 for survival to age 90 with cognitive impairment, and weaker associations in the range of 0.85-0.97 for survival to age 90 without cognitive impairment. The strongest associations were still observed for both outcomes for AgeAccelGrim2 and PCGrimAge but the odds ratios and 95% CIs were similar for survival to age 90 with and without cognitive impairment. The only clock that incorporated cognition in its development, Intrinsic Capacity (DNAmIC Age Accel), also was not more strongly associated with survival with preserved cognition (OR=1.16; 95% CI 1.08-1.24) than with survival without preserved cognition (OR=1.19; 95% CI 1.10-1.30).

**Figure 2:**
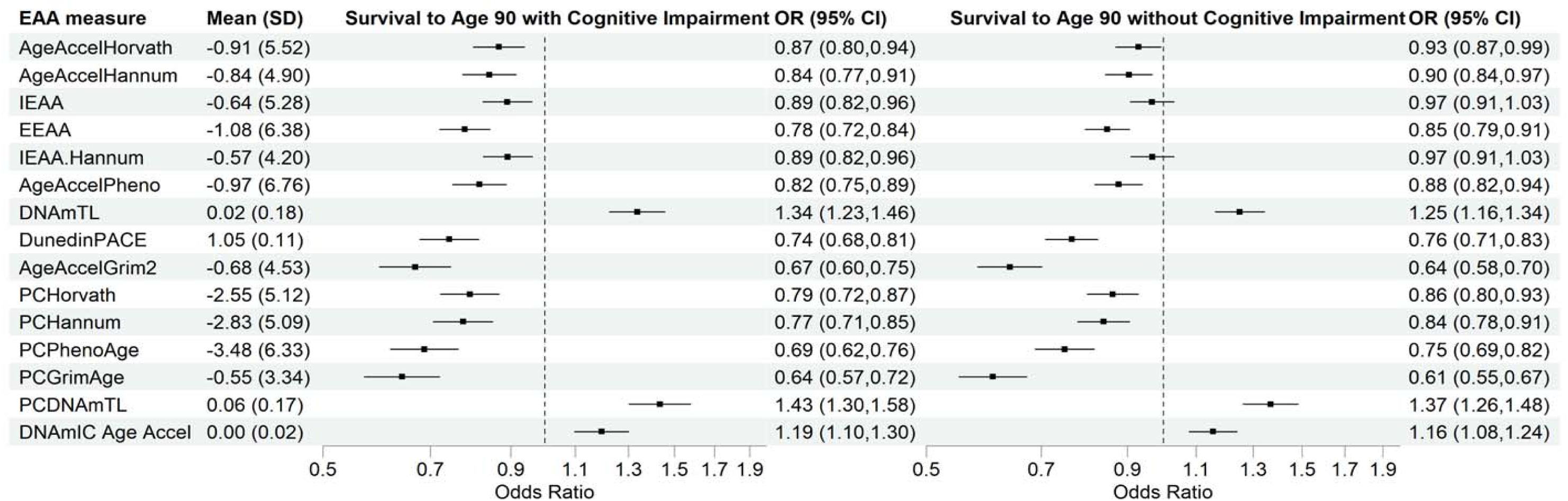
Odds ratios relating 15 epigenetic clocks to survival to age 90 with and without cognitive impairment among women enrolled in the Women’s Health Initiative Memory Study (WHIMS) who could have survived to age 90 (N=5,293) 1. The reference group for both survival groups was women who died before age 90 years (N = 2,611). 2. Odds ratios were adjusted for chronological age, race, ethnicity, hormone therapy treatment arm, education, smoking status, physical activity, body mass index, diabetes, cardiovascular disease, non-melanoma cancer, and white blood cell counts (CD8T, CD4T, natural killer cells, B cells, monocytes, and neutrophils). 3. Models with IEAA, EEAA, and IEAA.Hannum did not include white blood cell counts.

Analyses stratified by APOE ε2 or APOE ε4 carrier status showed no statistically significant interactions for survival to age 90 (Supplementary Tables 2 and 3), with the exception of APOE ε4 for DunedinPACE, which had an OR of 0.76 (95% CI 0.70-0.82) for non-carriers and 0.85 (95% CI 0.74-0.98) for carriers (p-interaction= 0.041). There were no significant interactions by APOE ε2 or APOE ε4 carrier status in analyses comparing survival to age 90 with and without preserved cognition vs. death before age 90 (Supplementary Tables 4 and 5). Results were unchanged when analyses were repeated including the women with missing race or ethnicity (data not shown).

## DISCUSSION

In this cohort study of 5,844 women, we found that all 15 epigenetic clocks examined were associated with exceptional longevity (survival to age 90 compared with death before that age). However, the magnitude of these associations differed markedly, with AgeAccelGrim2 and PCGrimAge having the strongest associations followed by PCPhenoAge and DunedinPACE; the odds of survival among these clocks ranged from 0.77-0.64. Overall, the first-generation clocks had the weakest associations, and the PC versions of the clocks had stronger associations than the original versions of the clocks. Unfortunately, none of the clocks were associated more strongly with survival to age 90 with preserved cognition vs. without preserved cognition compared to earlier death. Odds ratios were similar for both groups with overlapping confidence intervals, and the small differences observed were often in the direction of weaker associations with cognitively healthy longevity.

These results align well with previous meta-analyses of various epigenetic clocks in relation to total mortality. In a meta-analysis^13^ investigating five first-generation clocks using 13 population-based cohorts (n=13,089), modest summary hazard ratios for each year of age acceleration ranged from 1.01-1.03 with 95% confidence intervals, excluding one. Similarly, a subsequent systematic review and meta-analysis (11 studies, n=27,840) found hazard ratios for 5-year increments of EAA of 1.08 for AgeAccelHorvath and 1.15 for AgeAccelHannum. Most recently, 14 epigenetic clocks including second- and third-generation clocks were investigated in relation to 10-year mortality in the Generation Scotland cohort (n-18,859).^14^ Of the six clocks overlapping with this study, the hazard ratio for total mortality for a 1 standard deviation increase in EAA was strongest for GrimAge2 at 1.54 (95% CI 1.46-1.62). Hazard ratios were nearly as strong for DunedinPACE (1.41; 95% CI 1.32-1.50) and weaker but in the predicted direction for Hannum, Horvath, DNAmTL and PhenoAge. These associations with total mortality are predicting time to death irrespective of age at death. If a clock associates with time to death, it still may not differentiate those who will survive to the oldest ages vs. those who die at younger ages. Such studies are heavily influenced by early deaths and include people who were not followed long enough to reach the oldest age groups. The present study extends the mortality findings by observing a similar pattern with first-, second- and third-generation clocks with the outcome of survival to age 90 vs. death before age 90.

Due to limitations in cohort age ranges, follow-up lengths, and sample sizes, few studies have investigated epigenetic clocks in relation to survival to the oldest age groups. We previously reported in a separate subcohort of the Women’s Health Initiative (n=1830) on associations of four epigenetic clocks (AgeAccelHorvath, AgeAccelHannum, AgeAccelPheno, and AgeAccelGrim) in relation to survival to age 90 with intact mobility and cognition vs. death before age 90.^8^ All four measures of epigenetic age acceleration reduced the odds of survival to age 90 with intact mobility and cognitive function. AgeAccelPheno and AgeAccelGrim also reduced the odds of survival to age 90 without intact mobility and/or cognitive function, whereas the Hannum and Horvath clocks were not associated with survival to age 90 with impaired function. Odds ratios per standard deviation increase of AgeAccelPheno were stronger for healthy longevity (OR=0.60; 95% CI 0.50-0.72) than for longevity with impaired function (OR=0.74; 95% CI 0.62-0.88) in that study. For AgeAccelGrim, the odds ratios were similar for both survival groups vs. death before age 90. Results were similar when we examined survival to age 90 with impaired mobility alone, suggesting that impaired mobility at age 90 was driving the associations more than impaired cognition. Impaired mobility is a very strong risk factor for death.^15,16^

The present study was designed to focus specifically on longevity with preserved cognitive function to evaluate whether any of the most often studied clocks could differentiate survival with intact cognition vs. survival with impaired cognition in a larger study population followed for cognitive function and incident dementia. When intact cognition was required to define healthy longevity, none of the EAA measures were more associated with healthy survival than with cognitively impaired survival. While some studies have shown associations of EAA with various cognitive outcomes including cognitive functioning, mild cognitive impairment and dementia, a systematic review of 30 studies^17^ concluded that there was no consistent evidence that current measures of EAA are associated with these outcomes, and there was too much heterogeneity across studies to perform a meta-analysis. In a separate analysis of the WHIMS cohort,^11^ we found modest associations of AgeAccelGrim2 (HR= 1.11; 95% CI 1.02-1.20) and DunedinPACE (HR=1.07; 95% CI 1.01-1.15) with the combined outcome of incident MCI and dementia, but DunedinPACE was not associated with incident dementia alone. Dementia increases the risk of total mortality by almost six-fold,^18^ so it is highly likely to reduce survival to the oldest age groups in the majority of those afflicted.

Nonetheless, 16.4% of women in the present study survived to age 90 with impaired cognition defined as severe memory problems or dementia. These women may share aspects of slower biological aging related to physical aspects of their longevity but differ in ways related to their neurocognitive aging not measured by current epigenetic clocks. Indeed, the Intrinsic Capacity clock was the only EAA measure that was trained on a measure of cognitive function, the MMSE, as one of five health domains focused on physical, mental and sensory function. ^7^ It was associated with mortality in the Framingham Heart Study (HR=1.38) and with survival to age 90 in this study (OR=1.15), but did not differentiate survival to age 90 with and without cognitive function. Previous studies of adults aged 90 and older have shown that mixed pathologies underlying dementia are common in this age group, and also have shown substantial overlap in brain neuropathology findings between those with and without cognitive impairment.^19^ These complexities contribute to the challenges in developing epigenetic biomarkers that predict cognitively healthy longevity.

Strengths of this study include the large sample size of women followed through at least age 90, excellent retention and low loss to follow-up for survival status, rigorous surveillance of cognitive status for classification of severe cognitive impairments, the availability of rich covariate data, and the examination of first-, second- and third-generation epigenetic clocks. The WHIMS cohort is nested within the Women’s Health Initiative and restricted to women, which is a strength in the sense that women are more likely than their male counterparts to live into their 90s and have increased risk of developing dementia in their later years. It is also a limitation in the sense that these findings require replication in men. In a recent analysis of 14 clocks within the Generation Scotland cohort,^14^ the authors noted that clock-disease associations were generally consistent for men and women. We were unable to classify the cognitive status of 9.3% of women who did not have a cognitive assessment within three years of their 90^th^ birthday; however, the results for the clocks were very similar regardless of cognitive status at age 90. We examined 15 widely used epigenetic clocks and present odds ratios and associated 95% confidence limits to compare their magnitudes of association. Our purpose was not to establish the statistical superiority of any one clock over others. The consistency of these results with the literature on epigenetic clocks and mortality supports the reproducibility of these findings. It is not possible to infer causality of any epigenetic clock with the longevity outcomes, as experimental studies are needed to elucidate underlying causal mechanisms, an area that is moving slowly compared to the burgeoning examination of clock associations with many health outcomes.^20^

### Conclusion

An aspirational goal for epigenetic clocks as measures of biological aging is that they be capable of differentiating people who will have long survival with intact functioning vs. those who will die earlier or outlive their healthspans. Preserved cognitive function is especially salient, because older adults must retain this functionality to live independently. No clocks have been specifically developed focusing on this outcome, but using long-term longitudinal datasets following humans to their most advanced ages makes this possible, as illustrated here. If future clock development efforts are successful in showing reproducible and strong associations with healthy longevity, then the biomarkers can serve many purposes, including aiding in clinical decision-making about screening and life-prolonging treatments, individual life planning, testing of interventions designed to slow biological aging, and discovery of the underlying causality that produces these associations. The relatively strong associations with longevity for AgeAccelGrim2 observed here show promise, but it is critical to develop clocks that can differentiate long survival with and without intact cognitive function.

## Author Contributions

A.Z.L. and A.H.S. planned and designed the study. A.H.S. obtained funding. A.Z.L. performed literature review and drafted the manuscript. BZ conducted the statistical analysis and created the figures and tables. S.N. prepared the dataset and calculated the epigenetic clock data on participants, and A.X.M and C.N. led the processing of the epigenetic data for analysis. All authors contributed to data interpretation, manuscript review, and editing.

## Acknowledgements

We thank the WHI participants, staff, and investigators. The short list of WHI investigators can be found at: https://www-whi-org.s3.us-west-2.amazonaws.com/wp-content/uploads/WHI-Investigator-Short-List.pdf. The full list of WHI Investigators can be found at the following site: https://www.whi.org/doc/WHI-Investigator-Long-List.pdf

## Ethical Statement

Ethical approval for this study was obtained from the University of California San Diego Institutional Review Board.

## Consent

All participants provided written informed consent.

## Data Availability Statement

The data supporting the present study’s findings are available upon reasonable request and review of proposals submitted through the Women’s Health Initiative (WHI) study website at https://www.whi.org/propose-a-paper. Access is granted to investigators from recognized research institutions.

