## Supplemental Materials for "Epigenetic Clocks of Biological Aging and Cognitively Healthy Longevity: The Women’s Health Initiative Memory Study"

**Table of Contents**

**Supplementary Methods**

**Supplementary Table 1**: Baseline characteristics by quartiles of DunedinPACE in 5911 women enrolled in the Women’s Health Initiative Memory Study (WHIMS) who could have survived to age 90

**Supplementary Table 2**: Subgroup analysis of epigenetic clocks and survival to age 90 by APOE ε2

**Supplementary Table 3**: Subgroup analysis of epigenetic clocks and survival to age 90 by APOE ε4

**Supplementary Table 4**: Subgroup analysis of epigenetic clocks and survival to age 90 with and without cognitive impairment by APOE ε2

**Supplementary Table 5**: Subgroup analysis of epigenetic clocks and survival to age 90 with and without cognitive impairment by APOE ε4

**SUPPLEMENTARY METHODS**

**Analytic Sample**

Among 7,479 WHIMS participants, we excluded 240 with only one WHIMS cognitive assessment, 519 who were not dbGaP-eligible, and 304 with no baseline DNA or buffy coat available, leaving 6,416 participants whose baseline biospecimens were used for epigenetics measurement (Supplementary Figure 1). After quality control (see below), epigenetic clock data were available for 6,069 participants. We further limited analyses to women born on or before February 17, 1934, who were eligible, because of birth year, to survive to age 90 by the end of follow-up ending February 17, 2024, leaving 5,911 women in the study population. Sixty-seven women were excluded due to missing race or ethnicity, a possible confounder given its associations with both EAA and longevity, leaving 5,844 for the analysis of survival to age 90. An additional 551 women were excluded from the analysis of cognitively preserved longevity because their cognitive status at age 90 could not be classified.

**Methylation Quality Control**

Samples were assessed for quality using 17 control metrics from Illumina. Samples were removed if they did not meet Illumina’s recommended thresholds for each control metric (N=14 removed). Sex was determined by clustering samples on the average intensity values of CpG sites on the X and Y chromosomes.^1^ Samples that fell outside of the female cluster or had a mismatch between reported and detected sex were excluded (N=8 removed). Samples with mean bisulfite intensity values < 4,000 were excluded (N=1 removed). Detection p-values were calculated using out-of-band (OOB) probes.^2^ CpG measurements were set to missing if detection p > 0.05 or <= 3 detection beads (M=66,532 probes). CpG sites were removed if >5% of samples were missing data. Based on the remaining CpG sites, samples were excluded if >5% of CpG sites had missing methylation values (N=43 removed). In the remaining samples, OOB background correction, RELIC dye bias correction, and RCP probe type bias correction were applied using Enmix.^3^

**Sample concordance**

Concordance between samples was measured using the SNP fingerprinting probes built into the array. Illumina SNP probes were converted to genotype data. These genotypes were assessed for pairwise IBD in PLINK. If a sample had a 100% match with an unexpected sample, the pair was compared against genotype array data. Under the assumption that array genotypes represented the 'truth' dataset, the sample from the pair that did not match the array genotype was excluded.

**Relatedness**

Kinship was determined from genotype data using KING.^4^ For each pair of participants with estimated 3rd degree or closer relatedness, one was excluded, with the preference to retain cases. If case status matched, one of the pair was removed at random. In WHIMS, there were 7 first degree relative (parent-child, siblings) pairs in the entire data and 53 relative pairs were approximately 3^rd^ degree related (first cousins).

**WHI Protocol for Blood Collection and Processing**

<https://www.whi.org/doc/Vol-2-11-Blood-and-Urine-Collection-Processing-and-Shipment.pdf>

**Epigenetic Clocks**

AgeAccelHorvath, AgeAccelHannum, IEAA, EEAA, IEAA.Hannum, AgeAccelPheno, and DNAmTL were calculated with the online Horvath and Clock Foundation DNAm Age Calculator.^5^ IEAA and EEAA are first generation clocks.^5,6^ IEAA measures cell-intrinsic methylation changes, and EEAA tracks age-related changes in white blood cell-type composition. IEAA Hannum measures intrinsic epigenetic age acceleration according to Hannum’s clock.^7^ AgeAccelPheno is a second-generation clock capturing phenotypic age, consisting of age and 9 clinical biomarkers (e.g., creatinine); this clock was associated with lifespan and many indicators of healthspan.^8^ DNAmTL has been shown to outperform leukocyte telomere length (LTL) in detecting associations with age, sex, ethnicity, lifestyle factors, clinical biomarkers, mortality, and cardiovascular disease.^9^

DunedinPACE was developed using longitudinal data and captures the pace of aging across various organ systems (e.g., cardiovascular, immune, pulmonary); it has been shown to be associated with cognitive function, morbidity, and disability.^10^ DunedinPACE was calculated using R code available at GitHub from the Belsky Lab (<https://github.com/danbelsky/DunedinPACE>).

AgeAccelGrim2 is the age-adjusted residual of the GrimAge2 clock, which is a mortality estimator composed of DNAm-based surrogates of plasma proteins, a DNAm-based estimator of smoking pack-years, age, and sex.^11^

DNAm IC^12,13^ captures intrinsic capacity, a composite of physical function, cognitive function, depressive symptoms, visual acuity and hearing, and hand grip strength. DNAmIC^12^ was calculated as a weighted linear combination of CpG β-values using the published regression coefficients. We calculated DNAmIC Age Accel as age-residualized DNAmIC.^12^ Higher DNAmIC values indicate greater intrinsic capacity.

PC-based versions of clocks were calculated using principal components from CpG-level data as the input for prediction of biological age, which have been shown to improve reliability, minimize technical noise, and more strongly predict age-related phenotypes and mortality compared to the original versions of the clocks.^14^ PC-based clocks were calculated using R code available on GitHub from the Levine Lab (<https://github.com/MorganLevineLab/PC-Clocks>).

AgeAccelHorvath, AgeAccelHannum, IEAA, EEAA, IEAA.Hannum, AgeAccelPheno, AgeAccelGrim2, and DNAmTL were calculated with the online Horvath and Clock Foundation DNAm Age Calculator (<https://dnamage.clockfoundation.org/>). We calculated the PC epigenetic clocks using R code available at (<https://github.com/MorganLevineLab/PC-Clocks>) and subsequently regressed them on chronological age to compute their respective residual values. DunedinPACE was calculated using R code available at GitHub from the Belsky Lab (<https://github.com/danbelsky/DunedinPACE>). DNAm IC was calculated using R code available from the lead author of the paper (<https://github.com/msfuentealba/IC_clock>)

**Adjudication of Mild Cognitive Impairment and Dementia**

Briefly, participants completed the Modified Mini-Mental State Examination, with those scoring below specific cutpoints (<80 for those with ≤8 years of education and <88 for those with ≥9 years of education) completing a modified Consortium to Establish a Registry for Alzheimer’s Disease battery of neuropsychological tests and standardized tests. A physician performed a neuropsychiatric evaluation with optional computerized tomography (without contrast) and blood assay, and a friend or family member designated by the participant was interviewed regarding functional changes. Mild cognitive impairment diagnosis was based on Petersen’s criteria, and dementia diagnosis was based on *Diagnostic and Statistical Manual of Mental Disorders, Fourth Edition (DSM-IV)* criteria.^15,16^ All data were sent to the WHIMS Clinical Coordinating Center for review and central adjudication of final classification of cognitive status by an adjudication panel consisting of a neurologist, geriatric psychiatrist, and geropsychologist. WHIMS-ECHO used a common, validated protocol of telephone-based cognitive assessments and informant reviews, and a similar protocol to that of WHIMS for ascertainment and central adjudication of final diagnosis.^17^

Figure 1: Flow diagram describing the study population of WHI Memory Study women eligible to survive to age 90 with longevity and cognitive status outcomes

7,479 women in the Women’s Health Initiative Memory Study

6,416 women had DNA extracted from blood sample

Excluded:

- 240 with only one cognitive assessment
- 519 with did not consent to sharing DNA
- 304 with no baseline DNA or buffy coat available

Excluded:

- 347 excluded following QC
- 43 women with incomplete CpG site data

6,026 women with complete DNAm data

115 women born after Feb. 1934

5,911 women eligible to survive to age 90 as of February 2024

5,844 women with complete epigenetic clocks and survival status data

67 women excluded for missing race or ethnicity

5,293 women with information on cognitive status within 3 years of 90^th^ birth year or died before age 90

551 women excluded from analysis of cognitively intact survival for unknown cognitive status within three years of their 90^th^ birthday

Supplementary Table 1: Baseline characteristics by quartiles of DunedinPACE in 5911 women enrolled in the Women’s Health Initiative Memory Study (WHIMS) who could have survived to age 90

| **Characteristic** | **Overall N = 5,911** | **Q1: [0.511, 0.980) N = 1,478** | **Q2: [0.980, 1.049) N = 1,477** | **Q3: [1.049, 1.122) N = 1,478** | **Q4: [1.122, 1.642] N = 1,478** | **p-value** |
| --- | --- | --- | --- | --- | --- | --- |
| **Age, Mean (SD)** | 70.1 (3.8) | 70.0 (3.7) | 70.2 (3.7) | 70.4 (3.8) | 70.0 (3.8) | 0.02 |
| **Education, n (%)** |  |  |  |  |  | <0.001 |
| Less than high school equivalent | 412 (7.0%) | 68 (4.6%) | 92 (6.3%) | 112 (7.6%) | 140 (9.5%) |  |
| High school diploma or GED | 1,275 (21.7%) | 321 (21.8%) | 327 (22.2%) | 328 (22.3%) | 299 (20.3%) |  |
| Vocational, training school, or some college or associate | 2,347 (39.9%) | 535 (36.3%) | 573 (38.9%) | 613 (41.6%) | 626 (42.6%) |  |
| College graduate or higher | 1,855 (31.5%) | 550 (37.3%) | 480 (32.6%) | 419 (28.5%) | 406 (27.6%) |  |
| Missing | 22 | 4 | 5 | 6 | 7 |  |
| **Smoking status, n (%)** |  |  |  |  |  | <0.001 |
| Never smoked | 3,100 (53.2%) | 912 (62.6%) | 849 (57.9%) | 755 (52.1%) | 584 (40.3%) |  |
| Past smoker | 2,319 (39.8%) | 515 (35.3%) | 558 (38.1%) | 605 (41.7%) | 641 (44.2%) |  |
| Current smoker | 403 (6.9%) | 30 (2.1%) | 59 (4.0%) | 90 (6.2%) | 224 (15.5%) |  |
| Missing | 89 | 21 | 11 | 28 | 29 |  |
| **Race, n (%)** |  |  |  |  |  | <0.001 |
| American Indian or Alaskan Native | 13 (0.2%) | 2 (0.1%) | 4 (0.3%) | 2 (0.1%) | 5 (0.3%) |  |
| Asian | 103 (1.8%) | 15 (1.0%) | 20 (1.4%) | 41 (2.8%) | 27 (1.9%) |  |
| Native Hawaiian or other Pacific Islander | 7 (0.1%) | 0 (0.0%) | 3 (0.2%) | 1 (0.1%) | 3 (0.2%) |  |
| Black | 406 (6.9%) | 55 (3.7%) | 75 (5.1%) | 97 (6.6%) | 179 (12.3%) |  |
| White | 5,264 (89.8%) | 1,391 (94.4%) | 1,352 (92.1%) | 1,300 (88.7%) | 1,221 (84.0%) |  |
| More than one race | 67 (1.1%) | 10 (0.7%) | 14 (1.0%) | 25 (1.7%) | 18 (1.2%) |  |
| Missing | 51 | 5 | 9 | 12 | 25 |  |
| **Ethnicity, n (%)** |  |  |  |  |  | <0.001 |
| Not Hispanic or Latino | 5,739 (97.4%) | 1,457 (98.9%) | 1,440 (97.7%) | 1,433 (97.3%) | 1,409 (95.7%) |  |
| Hispanic or Latino | 153 (2.6%) | 16 (1.1%) | 34 (2.3%) | 40 (2.7%) | 63 (4.3%) |  |
| Missing | 19 | 5 | 3 | 5 | 6 |  |
| **APOE ε2 carrier status, n (%)** |  |  |  |  |  | 0.9 |
| Non-carrier | 4,028 (84.7%) | 1,071 (84.3%) | 1,035 (84.3%) | 1,005 (85.2%) | 917 (85.1%) |  |
| Carrier | 727 (15.3%) | 199 (15.7%) | 193 (15.7%) | 174 (14.8%) | 161 (14.9%) |  |
| Missing | 1,156 | 208 | 249 | 299 | 400 |  |
| **APOE ε4 carrier status, n (%)** |  |  |  |  |  | 0.6 |
| Non-carrier | 3,573 (75.1%) | 937 (73.8%) | 933 (76.0%) | 893 (75.7%) | 810 (75.1%) |  |
| Carrier | 1,182 (24.9%) | 333 (26.2%) | 295 (24.0%) | 286 (24.3%) | 268 (24.9%) |  |
| Missing | 1,156 | 208 | 249 | 299 | 400 |  |
| **Physical activity (MET-hours/week), Mean (SD)** | 11.3 (13.2) | 13.7 (14.3) | 11.7 (13.1) | 10.7 (12.5) | 9.1 (12.2) | <0.001 |
| Missing | 14 | 3 | 2 | 3 | 6 |  |
| **BMI, Mean (SD)** | 28.5 (5.7) | 26.8 (5.1) | 28.0 (5.5) | 28.8 (5.4) | 30.6 (6.0) | <0.001 |
| Missing | 32 | 10 | 7 | 5 | 10 |  |
| **Diabetes, n (%)** | 374 (6.3%) | 30 (2.0%) | 72 (4.9%) | 97 (6.6%) | 175 (11.9%) | <0.001 |
| Missing | 10 | 2 | 1 | 3 | 4 |  |
| **Cardiovascular disease, n (%)** | 270 (4.6%) | 42 (2.8%) | 49 (3.3%) | 67 (4.5%) | 112 (7.6%) | <0.001 |
| **Non-melanoma cancer, n (%)** | 622 (10.5%) | 180 (12.2%) | 136 (9.2%) | 140 (9.5%) | 166 (11.2%) | 0.023 |
| **Hormone therapy treatment arm, n (%)** |  |  |  |  |  | <0.001 |
| Estrogen-alone placebo | 1,140 (19.3%) | 235 (15.9%) | 257 (17.4%) | 307 (20.8%) | 341 (23.1%) |  |
| Estrogen-alone intervention | 1,124 (19.0%) | 243 (16.4%) | 286 (19.4%) | 290 (19.6%) | 305 (20.6%) |  |
| Estrogen+Progestin placebo | 1,855 (31.4%) | 516 (34.9%) | 465 (31.5%) | 436 (29.5%) | 438 (29.6%) |  |
| Estrogen+Progestin intervention | 1,792 (30.3%) | 484 (32.7%) | 469 (31.8%) | 445 (30.1%) | 394 (26.7%) |  |

Supplementary Table 2: Subgroup analysis of epigenetic clocks and survival to age 90 by APOE ε2

|  | Non-carriers Survived to Age 90 (N = 2166) | | | Carriers Survived to Age 90 (N = 440) | | |  |
| --- | --- | --- | --- | --- | --- | --- | --- |
| Epigenetic Clock | Mean (SD) | OR (95% CI) | P Value | Mean (SD) | OR (95% CI) | P Value | P-interaction |
| AgeAccelHorvath | -0.62 (5.48) | 0.88 (0.82, 0.94) | < 0.001 | -0.77 (5.67) | 1.01 (0.86, 1.19) | 0.920 | 0.194 |
| AgeAccelHannum | -0.68 (4.83) | 0.85 (0.80, 0.92) | < 0.001 | -0.86 (4.67) | 0.97 (0.82, 1.15) | 0.723 | 0.260 |
| IEAA | -0.44 (5.21) | 0.91 (0.85, 0.97) | 0.004 | -0.60 (5.40) | 1.01 (0.86, 1.18) | 0.944 | 0.256 |
| EEAA | -0.83 (6.26) | 0.80 (0.75, 0.86) | < 0.001 | -1.04 (6.07) | 0.90 (0.77, 1.06) | 0.212 | 0.226 |
| IEAA.Hannum | -0.43 (4.16) | 0.91 (0.85, 0.97) | 0.003 | -0.54 (4.01) | 0.97 (0.83, 1.14) | 0.697 | 0.517 |
| AgeAccelPheno | -0.76 (6.71) | 0.84 (0.78, 0.90) | < 0.001 | -1.20 (6.68) | 0.94 (0.79, 1.12) | 0.490 | 0.216 |
| DNAmTL | 0.01 (0.18) | 1.24 (1.16, 1.33) | < 0.001 | 0.01 (0.19) | 1.25 (1.05, 1.50) | 0.013 | 0.957 |
| DunedinPACE | 1.05 (0.11) | 0.79 (0.73, 0.85) | < 0.001 | 1.04 (0.10) | 0.75 (0.62, 0.90) | 0.003 | 0.647 |
| AgeAccelGrim2 | -0.82 (4.51) | 0.65 (0.60, 0.72) | < 0.001 | -1.09 (4.32) | 0.69 (0.56, 0.87) | 0.001 | 0.527 |
| PCHorvath | -2.14 (5.04) | 0.80 (0.74, 0.86) | < 0.001 | -2.52 (4.81) | 0.89 (0.74, 1.07) | 0.202 | 0.427 |
| PCHannum | -2.54 (5.03) | 0.78 (0.72, 0.84) | < 0.001 | -2.93 (4.86) | 0.90 (0.74, 1.08) | 0.257 | 0.272 |
| PCPhenoAge | -3.52 (6.31) | 0.72 (0.65, 0.78) | < 0.001 | -3.95 (6.19) | 0.77 (0.62, 0.95) | 0.017 | 0.608 |
| PCGrimAge | -0.56 (3.36) | 0.63 (0.57, 0.70) | < 0.001 | -0.74 (3.29) | 0.76 (0.60, 0.96) | 0.022 | 0.163 |
| PCDNAmTL | 0.05 (0.17) | 1.38 (1.27, 1.50) | < 0.001 | 0.06 (0.17) | 1.33 (1.09, 1.63) | 0.005 | 0.979 |
| DNAmIC Age Accel | 0.00 (0.02) | 1.18 (1.10, 1.27) | < 0.001 | 0.00 (0.02) | 1.14 (0.96, 1.35) | 0.134 | 0.757 |

1. The reference group was women who died before the age of 90 years (Non-carriers: N = 1860; Carriers: N = 287)
2. Model adjusted for chronological age, hormone therapy treatment arm, education, smoking status, physical activity, body mass index, diabetes, cardiovascular disease, non-melanoma cancer, and white blood cell counts (CD8T, CD4T, natural killer cells, B cells, monocytes, and neutrophils).
3. Models with IEAA, EEAA, and IEAA.Hannum did not include white blood cell counts.
4. P-values for interactions were derived from likelihood ratio tests comparing nested models with and without the interaction term between epigenetic age acceleration and race.

Supplementary Table 3: Subgroup analysis of epigenetic clocks and survival to age 90 by APOE ε4

|  | Non-carriers Survived to Age 90 (N = 2046) | | | Carriers Survived to Age 90 (N = 560) | | |  |
| --- | --- | --- | --- | --- | --- | --- | --- |
| Epigenetic Clock | Mean (SD) | OR (95% CI) | P Value | Mean (SD) | OR (95% CI) | P Value | P-interaction |
| AgeAccelHorvath | -0.67 (5.55) | 0.88 (0.82, 0.95) | < 0.001 | -0.57 (5.40) | 0.94 (0.83, 1.06) | 0.326 | 0.301 |
| AgeAccelHannum | -0.69 (4.83) | 0.86 (0.80, 0.93) | < 0.001 | -0.76 (4.72) | 0.89 (0.78, 1.01) | 0.082 | 0.578 |
| IEAA | -0.47 (5.29) | 0.90 (0.84, 0.97) | 0.004 | -0.44 (5.08) | 0.98 (0.87, 1.10) | 0.731 | 0.249 |
| EEAA | -0.86 (6.23) | 0.81 (0.76, 0.87) | < 0.001 | -0.89 (6.22) | 0.83 (0.73, 0.93) | 0.002 | 0.824 |
| IEAA.Hannum | -0.41 (4.17) | 0.89 (0.83, 0.96) | 0.002 | -0.56 (4.05) | 0.97 (0.86, 1.09) | 0.567 | 0.379 |
| AgeAccelPheno | -0.77 (6.75) | 0.85 (0.79, 0.92) | < 0.001 | -1.03 (6.54) | 0.83 (0.73, 0.94) | 0.005 | 0.844 |
| DNAmTL | 0.01 (0.18) | 1.23 (1.14, 1.33) | < 0.001 | 0.01 (0.18) | 1.30 (1.13, 1.49) | < 0.001 | 0.946 |
| DunedinPACE | 1.05 (0.11) | 0.76 (0.70, 0.82) | < 0.001 | 1.05 (0.11) | 0.85 (0.74, 0.98) | 0.021 | 0.041 |
| AgeAccelGrim2 | -0.88 (4.46) | 0.65 (0.59, 0.72) | < 0.001 | -0.80 (4.56) | 0.68 (0.57, 0.81) | < 0.001 | 0.273 |
| PCHorvath | -2.24 (4.97) | 0.80 (0.74, 0.87) | < 0.001 | -2.08 (5.11) | 0.84 (0.72, 0.96) | 0.013 | 0.429 |
| PCHannum | -2.60 (5.02) | 0.78 (0.72, 0.85) | < 0.001 | -2.59 (4.97) | 0.84 (0.72, 0.97) | 0.018 | 0.258 |
| PCPhenoAge | -3.58 (6.25) | 0.74 (0.67, 0.81) | < 0.001 | -3.60 (6.44) | 0.68 (0.58, 0.81) | < 0.001 | 0.779 |
| PCGrimAge | -0.61 (3.36) | 0.65 (0.59, 0.72) | < 0.001 | -0.54 (3.31) | 0.65 (0.53, 0.78) | < 0.001 | 0.626 |
| PCDNAmTL | 0.05 (0.17) | 1.35 (1.24, 1.47) | < 0.001 | 0.05 (0.17) | 1.48 (1.26, 1.73) | < 0.001 | 0.933 |
| DNAmIC Age Accel | 0.00 (0.02) | 1.19 (1.10, 1.28) | < 0.001 | 0.00 (0.02) | 1.14 (1.00, 1.31) | 0.054 | 0.364 |

1. The reference group was women who died before the age of 90 years (Non-carriers: N = 1525; Carriers: N = 622)
2. Model adjusted for chronological age, hormone therapy treatment arm, education, smoking status, physical activity, body mass index, diabetes, cardiovascular disease, non-melanoma cancer, and white blood cell counts (CD8T, CD4T, natural killer cells, B cells, monocytes, and neutrophils).
3. Models with IEAA, EEAA, and IEAA.Hannum did not include white blood cell counts.

P-values for interactions were derived from likelihood ratio tests comparing nested models with and without the interaction term between epigenetic age acceleration and race.

Supplementary Table 4: Subgroup analysis of epigenetic clocks and survival to age 90 with and without cognitive impairment by APOE ε2

|  | Non-carriers Survived to Age 90 | | | | | Carriers Survived to Age 90 | | | | |  |
| --- | --- | --- | --- | --- | --- | --- | --- | --- | --- | --- | --- |
|  |  | With Cognitive Impairment  (N = 642) | | Without Cognitive Impairment  (N = 1176) | |  | With Cognitive Impairment  (N = 116) | | Without Cognitive Impairment  (N = 252) | |  |
| EAA | Mean (SD) | OR (95% CI) | p | OR (95% CI) | p | Mean (SD) | OR (95% CI) | p | OR | p | p-interaction |
| AgeAccelHorvath | -0.62 (5.48) | 0.83 (0.76, 0.92) | < 0.001 | 0.92 (0.85, 1.00) | 0.040 | -0.77 (5.67) | 1.01 (0.79, 1.28) | 0.952 | 1.06 (0.87, 1.28) | 0.564 | 0.375 |
| AgeAccelHannum | -0.68 (4.83) | 0.83 (0.75, 0.92) | < 0.001 | 0.87 (0.80, 0.95) | 0.001 | -0.86 (4.67) | 0.90 (0.70, 1.15) | 0.405 | 1.05 (0.86, 1.28) | 0.634 | 0.345 |
| IEAA | -0.44 (5.21) | 0.86 (0.78, 0.94) | 0.001 | 0.96 (0.89, 1.03) | 0.255 | -0.60 (5.40) | 1.01 (0.80, 1.27) | 0.957 | 1.07 (0.89, 1.29) | 0.478 | 0.463 |
| EEAA | -0.83 (6.26) | 0.77 (0.70, 0.84) | < 0.001 | 0.83 (0.76, 0.89) | < 0.001 | -1.04 (6.07) | 0.85 (0.68, 1.07) | 0.164 | 0.99 (0.83, 1.19) | 0.940 | 0.246 |
| IEAA.Hannum | -0.43 (4.16) | 0.89 (0.81, 0.97) | 0.011 | 0.93 (0.87, 1.01) | 0.076 | -0.54 (4.01) | 0.90 (0.72, 1.13) | 0.375 | 1.06 (0.88, 1.27) | 0.539 | 0.562 |
| AgeAccelPheno | -0.76 (6.71) | 0.79 (0.72, 0.88) | < 0.001 | 0.85 (0.78, 0.93) | < 0.001 | -1.20 (6.68) | 0.99 (0.77, 1.27) | 0.934 | 0.94 (0.77, 1.14) | 0.515 | 0.242 |
| DNAmTL | 0.01 (0.18) | 1.31 (1.18, 1.45) | < 0.001 | 1.22 (1.12, 1.33) | < 0.001 | 0.01 (0.19) | 1.46 (1.12, 1.90) | 0.005 | 1.29 (1.05, 1.59) | 0.016 | 0.476 |
| DunedinPACE | 1.05 (0.11) | 0.76 (0.68, 0.85) | < 0.001 | 0.79 (0.72, 0.86) | < 0.001 | 1.04 (0.10) | 0.73 (0.55, 0.97) | 0.030 | 0.74 (0.59, 0.92) | 0.007 | 0.950 |
| AgeAccelGrim2 | -0.82 (4.51) | 0.65 (0.57, 0.75) | < 0.001 | 0.63 (0.56, 0.70) | < 0.001 | -1.09 (4.32) | 0.67 (0.48, 0.94) | 0.019 | 0.74 (0.57, 0.95) | 0.020 | 0.355 |
| PCHorvath | -2.14 (5.04) | 0.77 (0.69, 0.86) | < 0.001 | 0.83 (0.76, 0.91) | < 0.001 | -2.52 (4.81) | 0.75 (0.57, 0.98) | 0.036 | 0.95 (0.77, 1.16) | 0.589 | 0.151 |
| PCHannum | -2.54 (5.03) | 0.76 (0.68, 0.86) | < 0.001 | 0.81 (0.74, 0.88) | < 0.001 | -2.93 (4.86) | 0.73 (0.55, 0.96) | 0.025 | 0.95 (0.77, 1.18) | 0.661 | 0.153 |
| PCPhenoAge | -3.52 (6.31) | 0.68 (0.60, 0.78) | < 0.001 | 0.74 (0.66, 0.82) | < 0.001 | -3.95 (6.19) | 0.64 (0.46, 0.89) | 0.007 | 0.82 (0.64, 1.04) | 0.105 | 0.683 |
| PCGrimAge | -0.56 (3.36) | 0.64 (0.55, 0.73) | < 0.001 | 0.60 (0.53, 0.67) | < 0.001 | -0.74 (3.29) | 0.79 (0.56, 1.12) | 0.185 | 0.72 (0.55, 0.95) | 0.020 | 0.224 |
| PCDNAmTL | 0.05 (0.17) | 1.44 (1.28, 1.62) | < 0.001 | 1.39 (1.26, 1.53) | < 0.001 | 0.06 (0.17) | 1.47 (1.10, 1.98) | 0.010 | 1.30 (1.04, 1.64) | 0.023 | 0.220 |
| DNAmIC Age Accel | 0.00 (0.02) | 1.25 (1.13, 1.39) | < 0.001 | 1.19 (1.09, 1.29) | < 0.001 | 0.00 (0.02) | 1.23 (0.95, 1.59) | 0.111 | 1.09 (0.90, 1.32) | 0.381 | 0.719 |

1. The reference group was women who died before the age of 90 years (Non-carriers: N = 1860; Carriers: N = 287)
2. Model adjusted for chronological age, hormone therapy treatment arm, education, smoking status, physical activity, body mass index, diabetes, cardiovascular disease, non-melanoma cancer, and white blood cell counts (CD8T, CD4T, natural killer cells, B cells, monocytes, and neutrophils).
3. Models with IEAA, EEAA, and IEAA.Hannum did not include white blood cell counts.
4. P-values for interactions were derived from likelihood ratio tests comparing nested models with and without the interaction term between epigenetic age acceleration and race.

Supplementary Table 5: Subgroup analysis of epigenetic clocks and survival to age 90 with and without cognitive impairment by APOE ε4

|  | Non-carriers Survived to Age 90 | | | | | Carriers Survived to Age 90 | | | | |  |
| --- | --- | --- | --- | --- | --- | --- | --- | --- | --- | --- | --- |
|  |  | With Cognitive Impairment  (N = 529) | | Without Cognitive Impairment  (N = 1183) | |  | With Cognitive Impairment  (N = 229) | | Without Cognitive Impairment  (N = 245) | |  |
| EAA | Mean (SD) | OR (95% CI) | p | OR (95% CI) | p | Mean (SD) | OR (95% CI) | p | OR | p | p-interaction |
| AgeAccelHorvath | -0.67 (5.55) | 0.81 (0.73, 0.91) | < 0.001 | 0.93 (0.86, 1.01) | 0.088 | -0.57 (5.40) | 0.96 (0.82, 1.13) | 0.646 | 0.95 (0.81, 1.11) | 0.502 | 0.174 |
| AgeAccelHannum | -0.69 (4.83) | 0.83 (0.74, 0.93) | < 0.001 | 0.88 (0.81, 0.96) | 0.003 | -0.76 (4.72) | 0.88 (0.74, 1.05) | 0.154 | 0.95 (0.80, 1.12) | 0.514 | 0.603 |
| IEAA | -0.47 (5.29) | 0.83 (0.75, 0.92) | < 0.001 | 0.96 (0.88, 1.04) | 0.300 | -0.44 (5.08) | 1.00 (0.85, 1.17) | 0.990 | 0.99 (0.85, 1.16) | 0.938 | 0.158 |
| EEAA | -0.86 (6.23) | 0.77 (0.69, 0.85) | < 0.001 | 0.84 (0.78, 0.91) | < 0.001 | -0.89 (6.22) | 0.81 (0.69, 0.95) | 0.010 | 0.86 (0.74, 1.00) | 0.053 | 0.835 |
| IEAA.Hannum | -0.41 (4.17) | 0.86 (0.78, 0.96) | 0.005 | 0.92 (0.85, 1.00) | 0.041 | -0.56 (4.05) | 0.96 (0.82, 1.12) | 0.606 | 1.03 (0.88, 1.20) | 0.693 | 0.411 |
| AgeAccelPheno | -0.77 (6.75) | 0.81 (0.72, 0.91) | < 0.001 | 0.86 (0.79, 0.94) | < 0.001 | -1.03 (6.54) | 0.82 (0.69, 0.97) | 0.021 | 0.84 (0.71, 0.99) | 0.037 | 0.720 |
| DNAmTL | 0.01 (0.18) | 1.31 (1.17, 1.47) | < 0.001 | 1.22 (1.12, 1.33) | < 0.001 | 0.01 (0.18) | 1.43 (1.20, 1.72) | < 0.001 | 1.26 (1.06, 1.50) | 0.011 | 0.943 |
| DunedinPACE | 1.05 (0.11) | 0.72 (0.64, 0.82) | < 0.001 | 0.76 (0.69, 0.84) | < 0.001 | 1.05 (0.11) | 0.85 (0.71, 1.02) | 0.086 | 0.80 (0.66, 0.96) | 0.016 | 0.067 |
| AgeAccelGrim2 | -0.88 (4.46) | 0.67 (0.57, 0.78) | < 0.001 | 0.64 (0.57, 0.72) | < 0.001 | -0.80 (4.56) | 0.65 (0.52, 0.82) | < 0.001 | 0.65 (0.52, 0.82) | < 0.001 | 0.632 |
| PCHorvath | -2.24 (4.97) | 0.76 (0.68, 0.86) | < 0.001 | 0.83 (0.76, 0.91) | < 0.001 | -2.08 (5.11) | 0.76 (0.63, 0.92) | 0.006 | 0.92 (0.77, 1.09) | 0.334 | 0.551 |
| PCHannum | -2.60 (5.02) | 0.75 (0.66, 0.85) | < 0.001 | 0.81 (0.74, 0.89) | < 0.001 | -2.59 (4.97) | 0.77 (0.63, 0.94) | 0.011 | 0.91 (0.75, 1.09) | 0.308 | 0.429 |
| PCPhenoAge | -3.58 (6.25) | 0.70 (0.61, 0.81) | < 0.001 | 0.76 (0.68, 0.84) | < 0.001 | -3.60 (6.44) | 0.61 (0.48, 0.76) | < 0.001 | 0.73 (0.59, 0.90) | 0.004 | 0.938 |
| PCGrimAge | -0.61 (3.36) | 0.66 (0.56, 0.77) | < 0.001 | 0.63 (0.56, 0.72) | < 0.001 | -0.54 (3.31) | 0.65 (0.51, 0.84) | < 0.001 | 0.53 (0.41, 0.69) | < 0.001 | 0.414 |
| PCDNAmTL | 0.05 (0.17) | 1.43 (1.25, 1.62) | < 0.001 | 1.34 (1.22, 1.49) | < 0.001 | 0.05 (0.17) | 1.53 (1.24, 1.88) | < 0.001 | 1.48 (1.21, 1.82) | < 0.001 | 0.957 |
| DNAmIC Age Accel | 0.00 (0.02) | 1.25 (1.12, 1.40) | < 0.001 | 1.19 (1.09, 1.30) | < 0.001 | 0.00 (0.02) | 1.22 (1.03, 1.46) | 0.024 | 1.13 (0.95, 1.34) | 0.155 | 0.689 |

1. The reference group was women who died before the age of 90 years (Non-carriers: N = 1525; Carriers: N = 622)
2. Model adjusted for chronological age, hormone therapy treatment arm, education, smoking status, physical activity, body mass index, diabetes, cardiovascular disease, non-melanoma cancer, and white blood cell counts (CD8T, CD4T, natural killer cells, B cells, monocytes, and neutrophils).
3. Models with IEAA, EEAA, and IEAA.Hannum did not include white blood cell counts.
4. P-values for interactions were derived from likelihood ratio tests comparing nested models with and without the interaction term between epigenetic age acceleration and race.
